# Prospective Evaluation of Transpancreatic Sphincterotomy Comparing to Needle-knife Precut in Difficult Biliary Cannulation: Short-and Long-term Outcomes

**DOI:** 10.1101/2020.03.10.20032797

**Authors:** Fatema Tabak, Guo-Zhong Ji, Lin Miao

## Abstract

**Background/Aims:** Transpancreatic sphincterotomy (TPS) can be an alternative approach of biliary access in difficult cannulation cases. We aimed to prospectively evaluate the efficacy and safety of TPS compared to needle-knife precut (NKP), considering the late consequences of both techniques.

**Methods:** A total of 122 enrolled patients have been divided into three groups based on the applied secondary cannulation techniques. Selective cannulation success, ERCP procedure findings, and immediate adverse events were compared between groups. We investigated the long-term outcomes during six-month after the procedure.

**Results:** Successful selective cannulation was achieved in 92.9% with TPS similarly to other groups. The mean procedure time was shorter in the TPS group without significant difference. Using TPS did not affect the rate of post-ERCP pancreatitis (PEP) with less frequent post-ERCP bleeding and perforation after TPS compared to NKP, without significant difference. Patients who received TPS, NKP, or both had no symptoms related to papillary stenosis or chronic pancreatitis during the follow-up period.

**Conclusions:** Using TPS was useful to achieve success cannulation in difficult cases with an acceptable PEP rate. Furthermore, it was associated with reducing bleeding and perforation rates comparing with NKP and no differences related to the long term consequences within the follow-up period.

## INTRODUCTION

Endoscopic retrograde cholangiopancreatography (ERCP) has become a common therapeutic intervention for several pancreaticobiliary conditions. Selective cannulation of the common bile duct (CBD), which is the key to the successful biliary therapeutic procedure, could be achieved after a few attempts of standard cannulation methods in around 80% of the cases ^1^.

According to the European Society of Gastrointestinal Endoscopy (ESGE) guidelines, biliary cannulation is defined as difficult if cannulation lasts longer than five minutes, success requires more than five attempts, or the guidewire accidentally passes the pancreatic duct at least twice. Therefore, additional cannulation methods are often needed in difficult cannulation cases. Difficult cannulation is frequently reported as a risk factor for adverse events with a probability of failed biliary cannulation ranges from 5% to 18% of cases ^1–3^.

Different techniques are reported in the literature regarding cannulation of the papilla in the case of cannulation difficulty ^4–7^. Transpancreatic sphincterotomy (TPS) is a technique used for exposing the bile duct orifice by making an incision through the septum between biliary duct and pancreatic. It is used when attempts with initial methods have failed and usually performed only by endoscopists experienced in ERCP. TPS involves the placement of a papillotome in the pancreatic duct and performing sphincterotomy in the direction of the bile duct, then extending the sphincterotomy to cannulate the biliary duct ^1,8,9^.

ERCP is an invasive procedure with an overall adverse events rate of approximately 4–11%, and the most common one is post-ERCP pancreatitis (PEP) ^10–12^. Using advanced cannulation techniques in a native papilla is considered a risk factor of PEP, ESGE suggested prophylactic pancreatic stenting in patients receiving TPS to decrease this risk ^1,13–15^. Pancreatic duct stricture or chronic pancreatitis could be developed after pancreatic sphincterotomy; therefore, a long follow-up period is needed to track these adverse outcomes.

Several retrospective studies compared the effects of different cannulation techniques on ERCP outcomes ^4,16,17^. However, more prospective studies are needed to clarify the role of TPS in the case of difficult biliary access. This study aims to offer a new prospective evaluation for the differences in efficacy and adverse events rate between TPS and NKP cannulation, as well as to evaluate the impact of TPS in developing ductal stricture or chronic pancreatitis (CP) during the six-month follow-up period.

## MATERIALS AND METHODS

### Data Source and Participants

This study was conducted in our Institute of Digestive Endoscopy between July 2016 and January 2018. 972 Patients underwent ERCP during the study period, difficult cannulation was reported in 209 patients with native papilla, 122 of them underwent NKP, TPS, or both during the procedure. For the purpose of analysis, patients were divided into three groups based on the secondary cannulation technique. The first group included patients who received KNP; 80 patients of overall difficult cannulation cases, the second group included 28 patients who received TPS, and the third group included 14 patients who needed sequential NKP following unsuccessful TPS.

The study’s protocol was approved by our hospital’s institutional review board, and all patients gave their written informed consent to participate in ERCP. We collected the data using a designed form to record the clinical features of each patient. Data describing the patients’ characteristics, including demographics, indications, procedure details, and ERCP related adverse events, were analyzed.

We classified the duodenal papilla morphology into; periampullary diverticulum, small papilla, protruding papilla, and regular papilla.

### ERCP procedure

All ERCP procedures were performed by experienced endoscopists in our center. Patients underwent therapeutic ERCP using side-view duodenoscope following an overnight fast under conscious sedation. Patients were monitored continuously during the procedure using a pulse oximeter, electrocardiography monitoring, and an automatic blood pressure recording device and supplementary oxygen were provided when needed.

Initial biliary cannulation was routinely performed with the guidewire-assisted technique. Needle knife precut or transpancreatic sphincterotomy was performed when selective biliary access failed after 5 minutes standard cannulation attempts or more than two pancreatic cannulations. Additional needle-knife incision was applied when TPS failed to reach deep biliary cannulations. Since the guidewire was already in the pancreatic duct while performing TPS, prophylactic placement of pancreatic stent was applied to all patients with multiple pancreatic cannulations. The protective effect of the pancreatic stent has been strongly suggested by current guidelines ^1,2^. Besides, prophylactic octreotide dose was administered to all patients before ERCP during the entire study period, but the nonsteroid anti-inflammatory drug (NSAID) suppositories were not used. The standard process in our hospital requires that all the patients should be hospitalized three days after the procedure for observation and serum amylase testing after the procedure. All the discharged patients were informed to stay in contact for any delayed adverse events,

### Definitions

According to ESGE criteria ^1^, cannulation was considered difficult if either took more than 5 minutes, if it needed more than five cannulation attempts on the papilla, or if the pancreatic duct was cannulated more than twice.

All adverse events were defined according to published criteria. Post-ERCP pancreatitis was defined as new or worsened abdominal pain with an elevated amylase at least three times as the upper limit of the normal level, at more than 24h after ERCP, requiring admission or prolongation of planned hospitalization. Post-procedural bleeding evidenced by a drop-in hemoglobin >2g/dl. Perforation was diagnosed according to the imaging evidence of intraperitoneal or retroperitoneal leakage of contrast agent observed under radioscopy.

### Data Analysis

Differences among different patient were determined by using Fisher’s exact test for categorical variables, and non-categorical variables with the Mann–Whitney U test. Univariate regression was performed with significant variables (p<0.05). All statistical analyses were performed using SPSS software version 23 for Windows.

## RESULTS

### Study Population

From July 2016 to January 2018, there were 972 ERCPs performed in our Digestive Endoscopy Center. We excluded the patients with the previous sphincterotomy who underwent ERCP for follow up, stent removing, or taking a biopsy. Patients with altered anatomy (Billroth ll, Roux-en-y reconstruction, and duodenum stricture) were also excluded. A total of 209 patients with native papilla had difficult cannulation. In 87 of them, the guidewire-assisted cannulation was successful in achieving deep cannulation. Accordingly, only 122 patients who received other cannulation techniques were enrolled and analyzed, including demographics, indications, procedure details, and procedure-related adverse events (see Fig. 1).

**Fig. 1.**
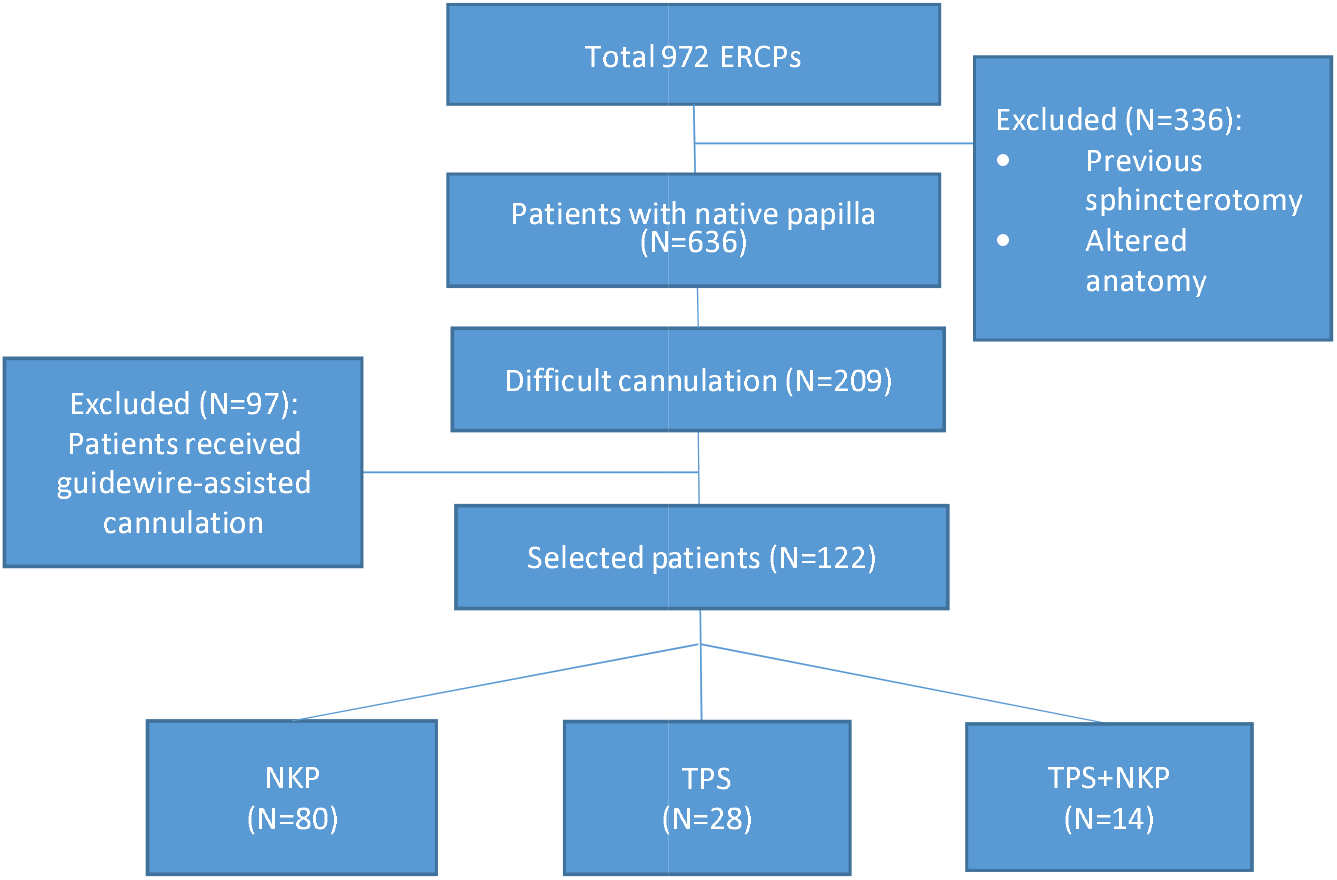
ERCP sample breakdown. ERCP, endoscopic retrograde cholangiopancreatography; TPS, transpancreatic sphincterotomy; NKP, needle-knife precut.

### Patient Characteristics

Based on the techniques used in difficult cannulation cases, 80 (65.5%) of patients were considered in the NKP group. The TPS group included 28 (22.9%) patients, and the third group who underwent TPS & NKP were 14 (11.4%) patients. The demographics and indications for ERCP are summarized in Table 1. There was no significant difference in patients’ gender in all groups, with a male percentage (52.5%, 67.9%, 42.9%, p=0.234). Using TPS was not associated with differences in the overall comorbidities (p=0.078), and the proportion of patients with Charlson Comorbidities Index (CCI) ≥2 was (31.2%% vs. 17.9%, 21.4%, p=0.343). The majority of the procedures were performed due to biliary stones in all groups; it was more common in the TPS group (66.2%,.78.6%, 50%, p=0.169), without any significant difference in the indication distribution or duodenal papilla morphology between groups.

**Table 1.** Characteristics’ summary and indications for ERCP.

| Parameter | Overall<br>(n=122) | Group A<br>NKP<br>(n=80) | Group B<br>TPS<br>(n=28) | Group C<br>NKP&TPS<br>(n=14) | p-value |
| --- | --- | --- | --- | --- | --- |
| **Age [mean ± standard deviation] (year)** | 65.29±16.33 | 68.36±14.95 | 58.43±18.40 | 61.43±15.66 | 0.237 |
| **Charlson score $\geq 2$** | 27% (33) | 31.2% (25) | 17.9% (5) | 21.4% (3) | 0.343 |
| **Male** | 54.9% (67) | 52.5% (42) | 67.9% (19) | 42.9% (6) | 0.234 |
| **Indications** |  |  |  |  |  |
| Biliary stones | 67.2% (82) | 66.2% (53) | 78.6% (22) | 50% (7) | 0.169 |
| Benign strictures | 21.3% (26) | 21.2% (17) | 17.9% (5) | 28.6% (4) | 0.726 |
| Cholangiocarcinoma | 4.9% (6) | 3.7% (3) | 3.6% (1) | 14.2% (2) | 0.227 |
| Cholangitis | 4.9% (6) | 6.2% (5) | 0% (0) | 7.1% (1) | 0.387 |
| Biliary pancreatitis | 4.1% (5) | 2.5% (2) | 10.7% (3) | 0% (0) | 0.120 |
| Chronic pancreatitis | 1.6% (2) | 0% (0) | 3.6% (1) | 7.1% (1) | 0.100 |
| Ampullary carcinoma | 3.3% (4) | 3.7% (3) | 3.6% (1) | 0% (0) | 0.764 |
| Pancreatic cancer | 9% (11) | 11.2% (9) | 0% (0) | 14.2% (2) | 0.154 |

### Success rate of biliary cannulation

Table 2 reports the details of cannulation and procedure after successful cannulation. The cannulation success rate did not differ among groups (P= 0.954). The overall success rate of biliary cannulation was 91.8% (112/122), including 91.2% (73/80) with NKP, 92.9% (26/28) with TPS, and 92.9% (13/14) with TPS & NKP. The three groups were comparable regarding the mean cannulation time (p =0.853) (see Fig. 2). However, using TPS was associated with shorter procedure duration (46.46±15.13) comparing with (53.54 ±27.55) in the NKP group and (56.43±31.03) in the TPS & NKP group, but did not reach to a significant difference. Significant differences have been noted in the rates of multiple pancreatic cannulations (31.2% vs. 92.9%, vs. 100%, p<0.001). After successful cannulation, no statistically significant differences were noted in the additional therapeutic interventions between the three groups.

**Table 2.**
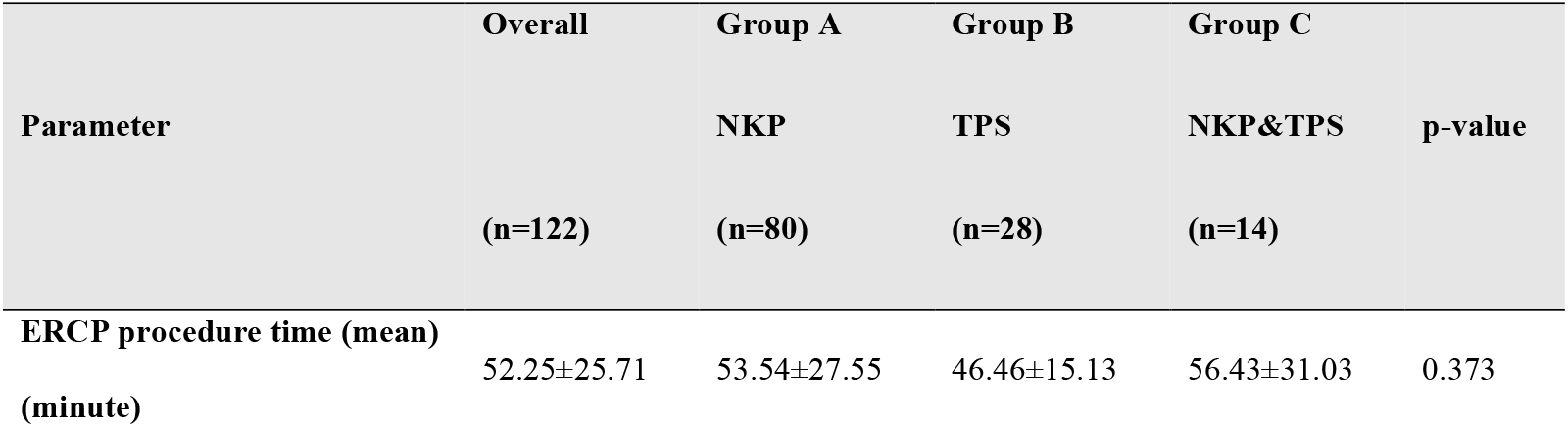
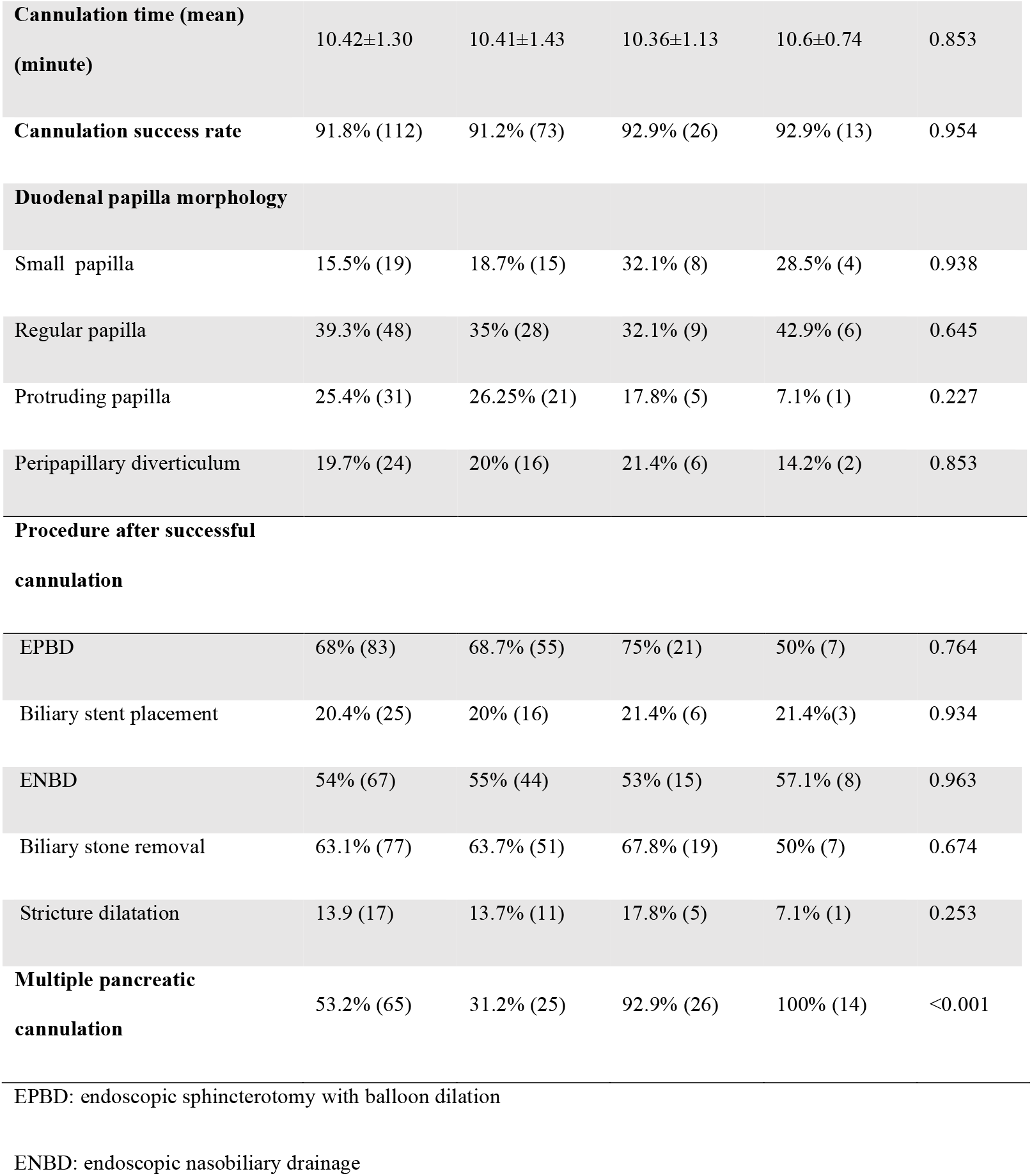
Details of cannulation and procedure after successful cannulation.

**Fig. 2.**
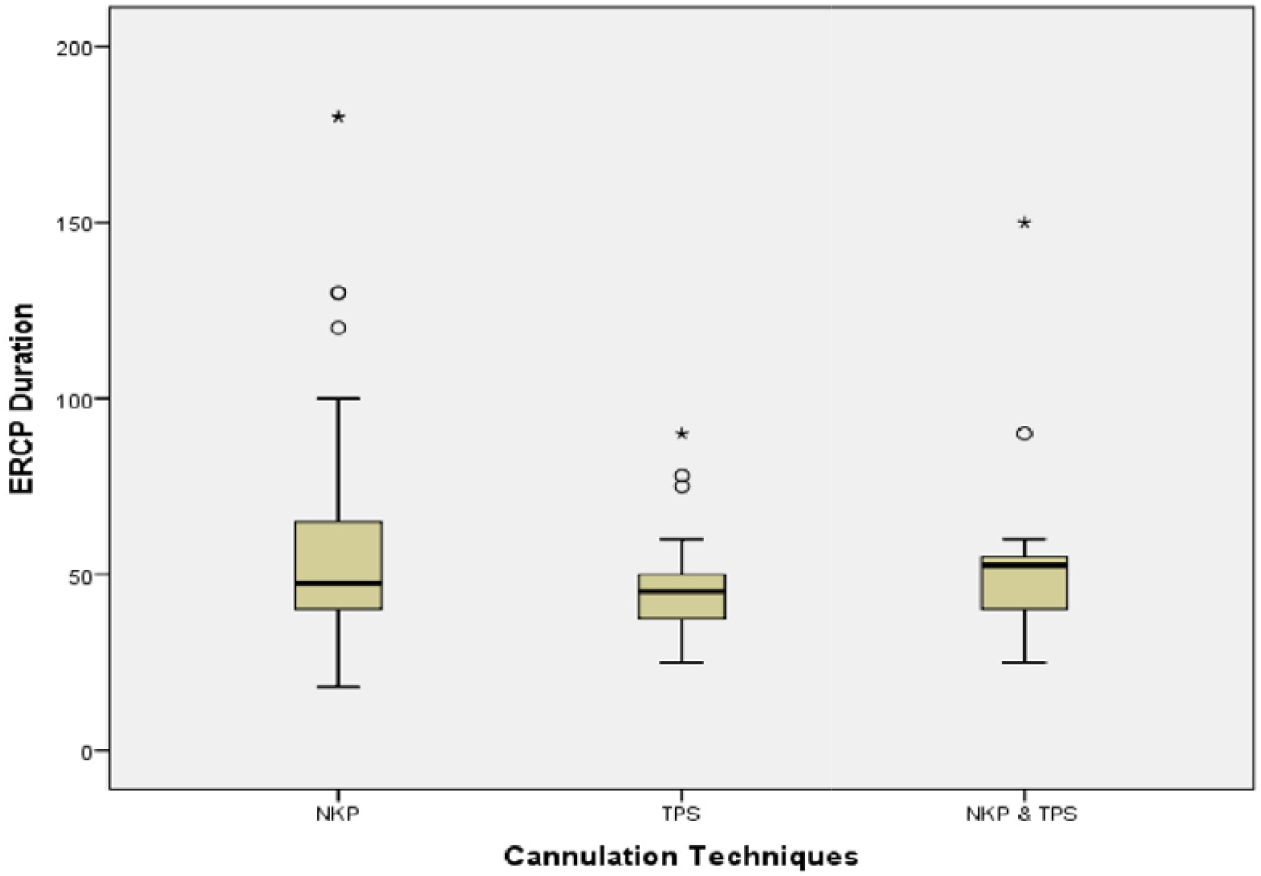
ERCP duration using cannulation techniques (NKP, TPS, and NKP&TPS) for the enrolled patients. The box is shorter for TPS than NKP&TPS, and NKP. The inner fences extend less for TPS compared to other groups. That is, the procedure time varies less for TPS than for NKP and NKP&TPS.

### Adverse events

The overall adverse events occurred in 12 patients (9.5%) (see Table 3), they were more frequent after NKP including 9 patients (11.2%), comparing with two patients (7.1%) in the TPS group, and one patient (7.1%) in the TPS & NKP group, albeit not reaching statistical significance (P = 0.495). The most common adverse event in all groups was post-ERCP pancreatitis, and no significant difference between groups regarding PEP frequency was seen in our study. There was a significantly higher rate of post-procedure hyperamylasemia (P = 0.021) in the TPS group (17.9%) and NKP following the TPS group (35.7%) when compared with NKP group (8.7%).

**Table 3.**
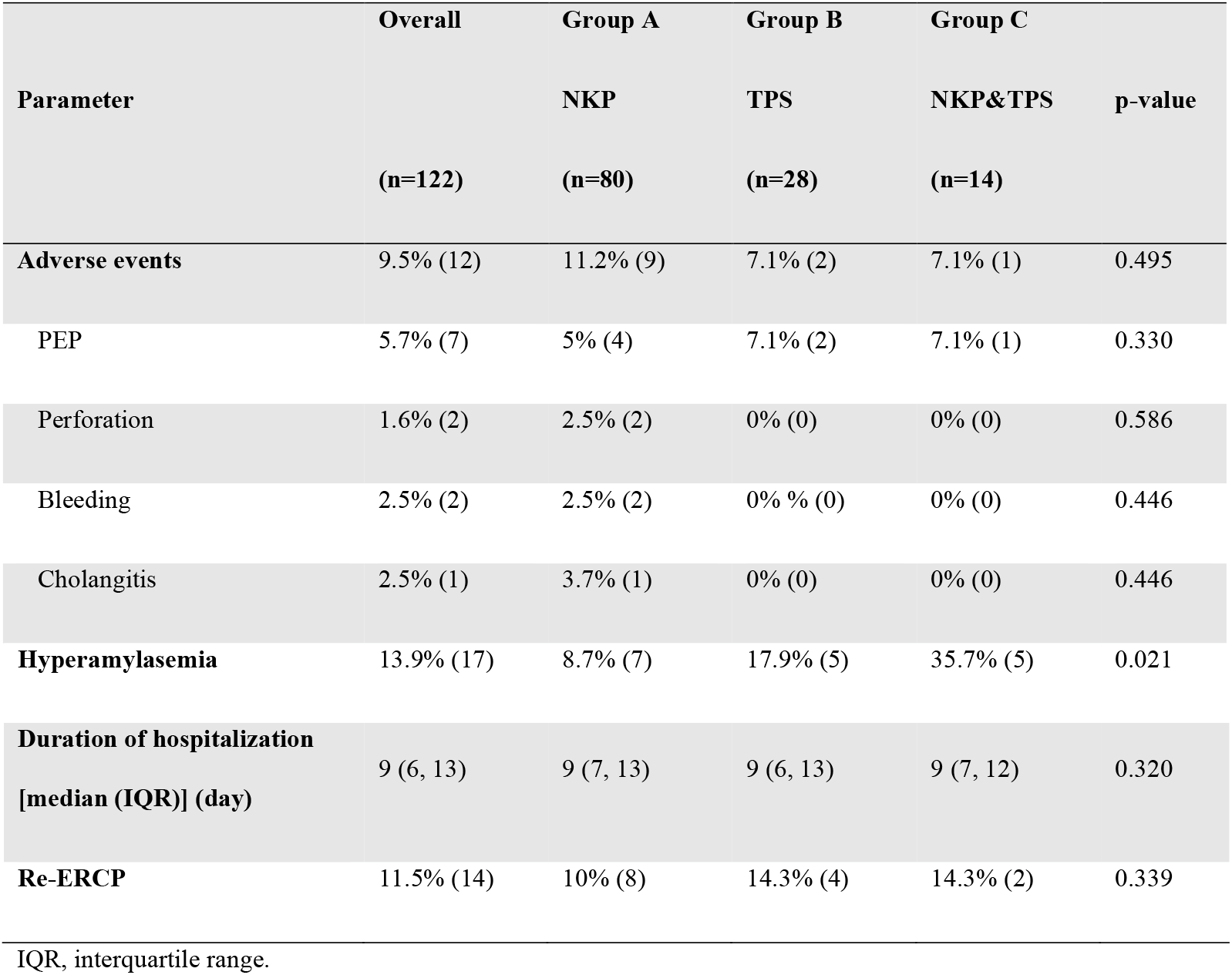
ERCP-related short-term and long-term outcomes.

The incidence of procedure-related bleeding and perforation did not differ significantly among groups, although the two bleeding and two perforation cases were in the NKP group. Both two bleeding cases were managed with endoscopic submucosal injection and/or placement of hemostatic clips, and blood transfusion when needed without surgical intervention. The two perforations cases were identified immediately at the time of ERCP and treated conservatively for around ten-days hospitalization with no requirement for surgical intervention. The patients’ hospitalization duration was similar in all groups (p=0.320).

All patients in the three groups were followed-up during six months after the procedure in the outpatient department to assess any long-term adverse events. During this period, patients in all three groups had no symptoms related to papillary stenosis, pancreatic strictures, or chronic pancreatitis. No cholangitis relapse was reported, two patients in the NKP group and one in the TPS group had biliary stones recurrence. 11.5% of patients need re-ERCP to complete stones clearance, exchange or remove stents with no differences among groups. Two patients in the NKP group died during the follow-up period due to tumor invasion.

## DISCUSSION

Success selective bile duct cannulation during ERCP can be achieved after a few attempts with standard guidewire-assisted cannulation in around 80% of cases. Recent guidelines recommended early applying advanced techniques in difficult canulation cases, including; needle-knife precut methods or transpancreatic sphincterotomy with the recommendation of prophylactic pancreatic stents (PPS) insertion in these cases ^1–3^. In our study, secondary cannulation performed with needle-knife fistulotomy technique in 80 patients (12.8%), transpancreatic sphincterotomy in 28 patients (4.45%), and an additional needle-knife incision was needed in 14 patients (2.24%) when TPS failed to reach deep biliary cannulations.

Regarding the efficacy of TPS, previous studies showed that TPS could be equally successful or even slightly better compared to other advanced cannulation methods in the case of difficult biliary access ^17–19^. The experience of the endoscopists, patients’ characteristics, and study design can be factors that reveal differences regarding cannulation success rate. The overall cannulation success rate of TPS in our study was favorably close to those reported in a relevant meta-analysis, which reported a success rate of around 90% by different study designs ^18,19^. Both TPS and KNP were effective in achieving successful biliary access without significant differences in the cannulation success rate and procedure duration. Even after a sequential NKP following TPS, the selective biliary cannulation was finally achieved in 92.9% of patients.

In the relation between TPS and adverse events, our study reported that patients who received NKP had a higher frequency of overall adverse events compared to the patients who received TPS or both techniques. However, we had not found any significant difference in overall adverse events, as many recent studies suggested ^16,17,19^.

Most studies showed that the PEP rate of TPS is similar to other advanced cannulation methods; even NKP could be better to avoid PEP ^18,20,21^. However, limited studies have prospectively compared PEP rate in TPS and NKP, in addition to lack of information on using uniform PEP preventive methods or pancreatic stent implantation in the TPS cases, which could make PEP rate in TPS even lower ^5,18^. The protective effect of pancreatic stent has been strongly suggested by ESGE, and its insertion is not problematic since the guidewire is already in the pancreatic duct while performing TPS ^1,18,20^.

In our study, we applied pancreatic stent in all patients undergoing TPS, in addition to the medical prevention of PEP, which was given to all patients before ERCP without using NSAID. Still, in this study, we noted an acceptable PEP rate of 7.1% in the two groups that used TPS without any significant difference comparing with the NKP group. In contrast, post-ERCP hyperamylasemia was significantly more common in the group that received both TPS and NKP compared with the other groups. It could be related to the multi pancreatic guidewire insertion and direct trauma to the pancreatic orifice because of several cannulation attempts using both techniques; even it does not make a statistically significant difference in the pancreatitis rate.

As stated in a recent meta-analysis, the bleeding rate of TPS was in the range of (2–4%), which was accepted compared to the approximate rate of (4%) for needle-knife precut techniques ^18^. The perforation rate of TPS was remarkably low compared with other techniques ^16,18^. In our study, no post-ERCP bleeding or perforation occurred after applying TPS. That could be related to the wire-assisted method of TPS, which gives better control of the cut than the freehand precut technique, which had acceptable bleeding and perforation rate.

Only a few prospective studies have evaluated the late adverse events of TPS ^9,22^. Our follow-up found no patient developed papillary stenosis, ductal strictures, or chronic pancreatitis during six months after the procedure and without any significant difference related to recurrent biliary stones or cholangitis among the three groups. Our results confirmed that TPS is an efficient method to achieve biliary cannulation in difficult cases and safe regarding early and late adverse events. Therefore, similarly to ESGE guidelines, we suggest early applying of TPS in difficult cannulation cases.

The main strength of this study is its prospective nature, using uniform PEP preventive methods in all TPS cases. in addition to six months follow-up period. There are few limitations in this study; the cases were collected at a single center with a relatively small sample included in the TPS & NKP group, so further prospective studies with a more extended follow-up period are still required to evaluate the long-term risks of TPS.

In conclusion, this study has shown that TPS is a useful rescue method in difficult biliary cannulation when performed by experienced endoscopists. It is well tolerated and having an acceptable adverse events rate, with a similar rate of post ERCP pancreatitis in patients receiving NKP. Also, sequential NKP following unsuccessful TPS appeared to be effective without increasing related adverse events rate. Additionally, our study demonstrated that no significant differences were noted in TPS patients and other groups regarding papillary stenosis, ductal strictures, or development of CP during the follow-up period.

## Data Availability

The datasets used and analyzed during the current study are available from the corresponding author on reasonable request.

## Conflicts of interests

The authors declare no conflict of interest.

## Notes

### Competing Interest Statement

The authors have declared no competing interest.

### Clinical Trial

NCT03771547

### Funding Statement

This research received no external funding.

